# The Moderating Role of Parental Sleep Knowledge on Children with Developmental Disabilities and Their Parents’ Sleep

**DOI:** 10.1101/2020.12.02.20242610

**Authors:** Elizabeth J. Halstead, Alexandra Jones, Gianluca Esposito, Dagmara Dimitriou

**Affiliations:** Sleep Education and Research Laboratory (SERL), UCL Institute of Education, London, WC1H 0AA United Kingdom; Social and Affective Neuroscience Lab, Psychology Program - SSS, Nanyang Technological University,Singapore; Lee Kong Chian School of Medicine, Nanyang Technological University, Singapore; Affiliative Behaviour and Physiology Lab, Department of Psychology and Cognitive Science, University of Trento, Italy

**Keywords:** Sleep, Children, Intellectual Disabilities, ASD, Developmental Disabilities, Sleep Knowledge, Parent Sleep Impairment

## Abstract

**Background:** Children with intellectual and developmental difficulties often experience 2 sleep problems, which in turn may impact parental sleep patterns. This study explored the role of parental sleep knowledge as a moderator on the relationship between child sleep and parental sleep impairment.

**Methods:** 582 parents or caregivers (92.6% mothers) of children with different developmental disabilities (Age *M* = 9.34, 29.5 % females) such as Down’s syndrome, participated in an online survey. Multiple regression analysis was conducted.

**Results:** Parental sleep knowledge of child sleep was a moderating variable in the relationship between child sleep nocturnal duration and parental sleep impairment. Although overall, sleep knowledge was high in this sample, two specific knowledge gaps were identified namely child sleep duration requirements, and the recognition of signs of a well-rested child.

**Conclusion:** This study has provided evidence that increased parental sleep knowledge can positively impact both child and parental sleep outcomes.

## 1. Introduction

Intellectual and developmental disabilities (IDD), such as Cerebral Palsy (CP), Down’s syndrome (DS) or Autism Spectrum Disorders (ASD), are diagnosed when a child presents with severe impairments of their cognitive, social and adaptive functioning [1]. Children with intellectual and developmental disabilities (IDD) experience more severe and longer lasting sleep problems than typically developing (TD) children [2]. Previous research reports that between 40-75% of children with IDD have problems with initiating and maintaining their sleep [3,4]. In addition, sleep disordered breathing, parasomnias (e.g. night terrors, sleep walking), and restless legs syndrome and/or periodic limb movement syndrome are often reported in children with IDD [5]. There is an established relationship between mental health and sleep in this population. For example, Owens and Palermo [6] found that 75% of children with major depressive disorder had insomnia symptoms. In addition, children with ASD are more likely to experience sleep problems if they have comorbid psychiatric issues, such as anxiety or depression [7]. Children with IDD experiencing sleep problems often require considerable care throughout the night, adversely impacting parental sleep [8]; and predicting parents’ sleep [9,10]. Approximately half of all mothers of children with IDD experience interrupted sleep due to caregiving needs of their children [11], with many waking more than twice in a night with less than a total of 7 hours of sleep [12]. Consistent with these studies, Gallagher *et al*. [13] found that 72% of caregivers to children with IDD, considered themselves to be sleep deprived [13]. Goldman *et al*. [14] studied 15 children with Angelman syndrome and found poor child sleep had a strong association with poor parental sleep, and increased parental stress. Impaired sleep in parents has also been associated with both physical and mental health, including an increased risk of cardiovascular and metabolic disease [15,16] and a weakened immune system [17]. Chronically sleep deprived parents frequently suffer from increased rates of psychiatric disorders and poorer psychological well-being [9,18]. Specifically, poor maternal sleep has been found to predict maternal depression [10]. Although the impact of poor sleep on mental health is evident in parents of children with IDD, Lee [19] reported that in mothers of children with IDD who experienced elevated stress, sleep problems, and depressive thoughts, a bidirectional relationship was present between depressive symptoms and poor sleep in mothers of children with IDD, highlighting the reciprocal influences at play. In addition, multiple studies have found parental stress to be the most robust predictor of inadequate parental sleep quality [13,20–22]. Poor maternal mental health along with family disruption, and deficient parent-child relationships have also been associated with children’s sleep disorders [8]. However, lack of research on the causal pathways and contributing factors to the relationship between child and parent sleep is evident [12,19].

If sleep problems in children with IDD and their parents are to be abated, further understanding of any influencing variables on the relationship between child and parent sleep is required. Research has suggested that sleep education may be an important factor for improving both child sleep and parental sleep. Sleep education emphasises the importance of bedtime routines, sleep duration requirements, minimisation of parent interactions during the night, and essential daytime, evening and bedtime habits which assist in the promotion of sleep [23]. A lack of sleep education specific to parents of children with IDD has been identified [24,25]. Parents often assume their child’s sleep issue is intrinsic to their IDD, thus underestimating the severity or even existence of their child’s sleep problems. [26–28]. This often means that parents do not seek treatment for their child’s problematic sleep [28]. One study found that 40% of parents reported their child with IDD waking regularly during the night but did not consider this to be a sleep problem [28]. Conversely, Ra *et al*. [29] found that the majority of the 58 parents correctly identified their child with IDD as having a severe sleep issue, however, these participants held their child’s IDD responsible for these sleep issues. Children are dependent on their parents possessing knowledge of sleep to ensure their sleep difficulties are appropriately treated and their sleep needs are met [30]. This is particularly pertinent for non-verbal children with IDD who may be unable to communicate their sleep related disturbances [31]. Although there are considerations for the delivery of this sleep education and best practices, due to the specific needs and individual circumstances of children with IDD and their families, [32], it is important to provide parents with the tools to implement positive sleep practices via sleep education. This may better equip parents to manage both parental and child sleep problems in IDD populations [33].

This study aims to partially replicate previous research in establishing if a relationship exists firstly between child sleep problems and parental sleep impairment and secondly, between nocturnal sleep duration of the child with IDD and parental sleep impairment. In addition, this study aims to establish the role of parental sleep knowledge as a potential moderator between these two relationships. Should a moderating relationship exist between child sleep problems or nocturnal sleep duration and parental sleep impairment, providing parents with appropriate education and assisting them to implement child behavioural change could offer important benefits to families experiencing sleep problems. To further explore parental sleep knowledge in this population, we report the percentage of correct answers to each of the sleep knowledge questions to identify specific knowledge gaps.

## 2. Materials and Methods

### 2.1. Participants

582 parents or primary caregivers of children with a diagnosed IDD completed an online survey. These included 539 mothers, 20 fathers, 18 adoptive mothers, 2 stepmothers, 1 aunt, and 2 grandmothers of a child with an IDD. 574 of the participants were primary carers for their child and 8 were secondary carers. All parents/caregivers reported that they were living with their child at the time of the completing the questionnaire. Children were aged between 2 and 17 years old (*M* = 9.35; *SD* = 3.34); 410 were males and 172 females. Primary diagnosis of children are included in Table 1. All children were considered to have one primary diagnosis, secondary conditions and diagnosis are included in Table 2. Average sleep hours of child primary diagnosis groups are reported in Table 3.

**Table 1.**

| Primary Diagnosis | N | Percent | Boys (n) | Girls (n) |
| --- | --- | --- | --- | --- |
| ASD | 429 | 73.7% | 320 | 109 |
| IDD | 78 | 12.4% | 41 | 37 |
| ADHD | 32 | 5.5% | 22 | 10 |
| Down's Syndrome | 17 | 2.9% | 8 | 9 |
| Cerebral Palsy | 8 | 1.4% | 8 | 0 |
| FASD | 7 | 1.2% | 4 | 3 |
| SEMH** | 6 | 1% | 4 | 2 |
| Sensory Processing Disorder | 5 | 0.9% | 3 | 2 |
| Total | 582 | 100% | 410 | 172 |

Abbreviations include: Autism Spectrum Disorder (ASD), Intellectual and Developmental Disabilities (IDD), Attention Deficit Hyperactivity Disorder (ADHD), Fetal Alcohol Syndrome Disorder (FASD), Social, Emotional & Mental Health (SEMH), please note this term is used in the UK Special Educational Need and Disabilities (SEND) Code of Practice (2014). \*Examples Include; Global Developmental Delay, Cornelia de Lange Syndrome and 22q11.2 deletion syndrome. \*\* Examples include: Anxiety, Complex Bereavement Disorder

**Table 2.**
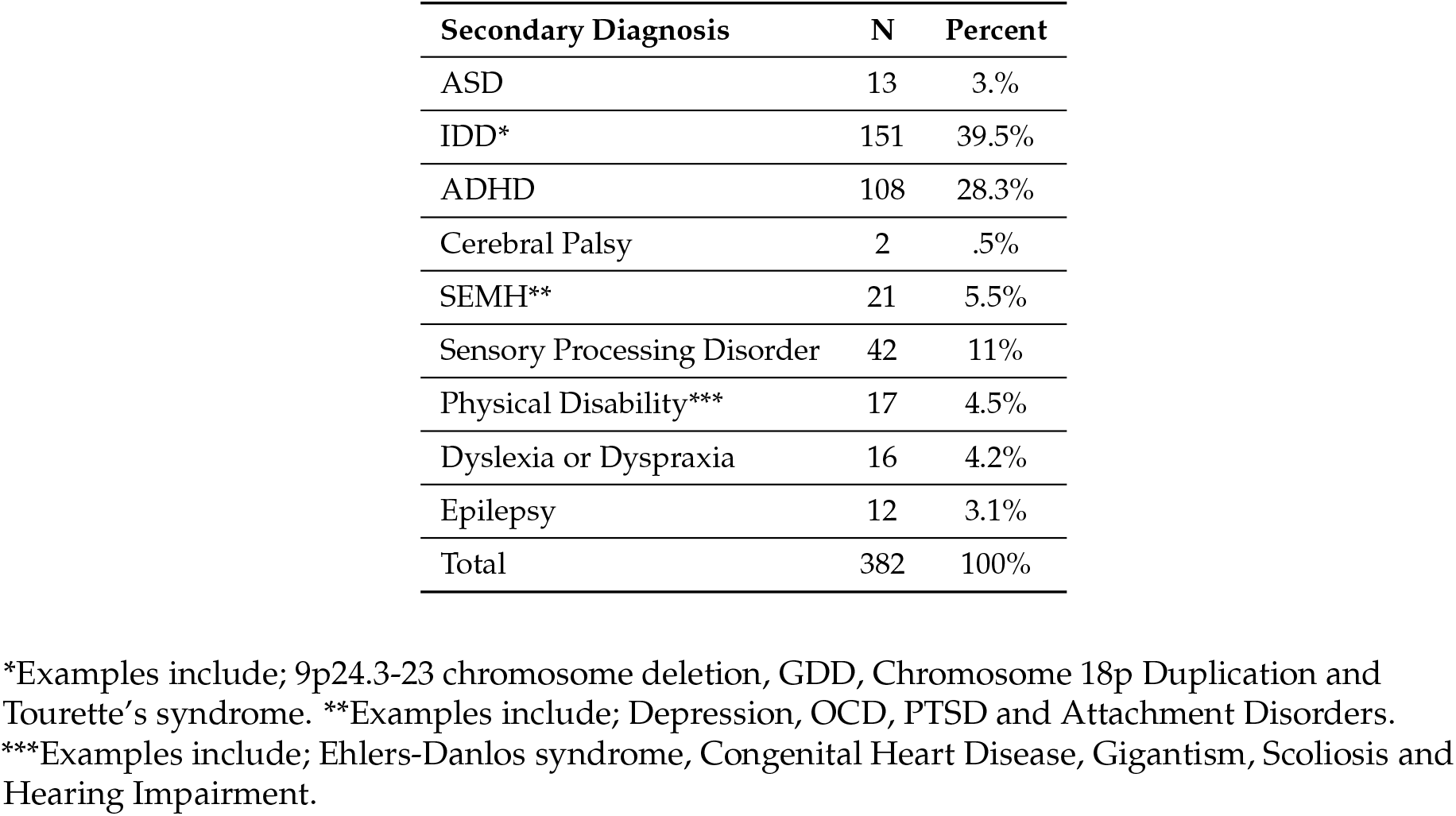

**Table 3.**

| Av. child sleep hours (per night) | M | SD |
| --- | --- | --- |
| ASD | 6.16 | 2.12 |
| IDD | 7.01 | 2.45 |
| ADHD | 6.03 | 2.37 |
| Down's Syndrome | 7.53 | 2.58 |
| Cerebral Palsy | 6.38 | 2.58 |
| FASD | 6.15 | 2.17 |
| SEMH** | 7.58 | 3.11 |
| Sensory Processing Disorder | 7.10 | 2.13 |

### 2.2. Procedure

This study was approved by the UCL institutional research review board in the UK (UCL IRB REC 1303). Participants were recruited to complete an online questionnaire through a multipoint recruitment method, which included emailing IDD parent support groups and UK non-governmental organizations (NGOs). Online recruitment via social media (Twitter and Facebook) was ongoing throughout the recruitment period. The REDCAP only survey system was used as the online system to collect the data. As all parents or caregivers completed all questions, there was no missing data in this dataset. Due to the nature of the recruitment methods, we were unable to determine the overall response rate for this survey.

### 2.3. Measures

Three measures plus a demographic questionnaire were utilised in the present study. All measures were completed by the parent or primary caregiver of the child with IDD.

#### 2.3.1. Demographic questionnaire

Demographic information about the child with IDD and their parent or guardian was gathered using a questionnaire developed by the researchers (see participants). The questionnaire included child age, gender, primary diagnosis, additional diagnosis or conditions, and the respondent’s relationship to the child.

#### 2.3.2. Child Sleep Problems and Nocturnal Sleep Duration

An abbreviated version of the Children’s Sleep Habit’s Questionnaire (CSHQ-A) was used to assess children’s sleep [34]. This 22-item version was modified from The CSHQ (Owens et al., 2000) and captured children’s sleep patterns (e.g. bed time, wake up time), behaviours (e.g. child falls asleep within 20 minutes after going to bed) and disturbances over a week period. Answers to questions were gathered on Likert scale of 1 (always) to 5 (never). Twelve questions were reversed scored to ensure higher scores were consistently attributed to sleep disturbance behaviours, e.g. ‘child awakens during the night and is sweating or screaming or inconsolable.’ The total score for each item was summed to provide an overall child sleep habits score, in addition to a total child nocturnal sleep duration. The present questionnaire produced a Cronbach’s Alpha score of .73.

#### 2.3.3. Parental Sleep Impairment

The PROMIS Item Bank v.1.0 – Sleep-Related Impairment [35] was used to measure the extent of sleep disturbances experienced by parents in the last week. The 16-item questionnaire assessed parents’ or carers’ bedtime routines, sleep hygiene practices, daytime behaviours and emotions relating to sleep in the past week (e.g. ‘I believe that stress disturbed my sleep,’). Responses were on a Likert scale of 1 (never) to 5 (always). The total score for each item was summed to provide an overall parental sleep impairment score. A score of above 50 was considered abnormal and reflected increased sleep impairment. In the present study, the Cronbach’s alpha score for this measurement was .73.

#### 2.3.4. Parental Sleep Knowledge (moderating variable)

The Sleep Knowledge Questionnaire devised by Owens *et al*. [36] was used to assess parental knowledge and beliefs about child sleep. Eight statements such as “snoring indicates a child is sleeping well,” were presented and parents selected whether the statements were true or false Owens *et al*. [36]. Correct answers were scored as 1 and incorrect answers were scored as 0. The higher the total score, the more knowledgeable parents were regarding positive sleep hygiene practices.

### 2.4. Data Analysis

Tests of normality were conducted on the four variables: child nocturnal sleep duration, child sleep problems, parent sleep related impairment, and parental sleep knowledge and Spearman’s Rho correlation analysis was used to explore the association between these variables. The Kruskal-Wallis one-way analysis of variance, correlations, and t-tests were conducted to explore the relationship between child demographics (age, sex, primary diagnosis) and the four variables. Significant relationships were added to the regression model as control factors. “PROCESS” custom dialog box [37] installed into SPSS® analytics software version 26.0 (IBM Corporation, Armonk NY, USA) for Windows® was used for the moderated multiple regression 126 analysis. To examine parental knowledge as a potential moderator, two multiple regression analyses were conducted to predict parental sleep related impairment. In the first analysis (model 1), child sleep problems was entered as a predictor. Parental sleep knowledge was also entered as a predictor variable along with the interaction term between child sleep problems and parental sleep knowledge. In the second analysis (model 2) child nocturnal sleep duration was entered as a predictor. Parental sleep knowledge was also entered as a predictor variable along with the interaction term between child nocturnal sleep duration and parental sleep knowledge. Child nocturnal sleep duration, child sleep problems, and parental knowledge variables were automatically mean-centred when using the PROCESS dialog box (the variable mean is subtracted from every value of the variable). Multicollinearity issues between variables were checked using the Variance Inflation Factor (VIF) and the variables showed no multicollinearity problems (all values < 10, average > 1, tolerance > 0.1; [38]). To further assess parental knowledge, the percentages of each question answered correctly are reported.

## 3. Results

Child nocturnal sleep duration was significantly correlated with child sleep problems, parental sleep related impairment, and parental knowledge. As child nocturnal sleep duration increased, child sleep problems decreased, parental sleep related impairments decreased and parental knowledge increased (*ps* < .001). Child nocturnal sleep duration accounted for a small variance of 3.4% in parent sleep related impairment score. Child sleep problems was significantly correlated with parental sleep related impairment. As child sleep problems increased, parental sleep impairment also increased (*p* < .001). Child age was found to negatively correlate to child total nocturnal sleep duration (*p* = .001) and overall child sleep problems (*p* < .001). As child’s age increased, the total nocturnal sleep duration per night and child sleep problems decreased. See Table 4 for correlation coefficients.

**Table 4.** Inter-correlations coefficients for the variables: Child Age, Child Nocturnal Sleep Duration, Parent Sleep Related Impairment and Parental Sleep Knowledge.

| Variables | 1 | 2 | 3 | 4 | 5 |
| --- | --- | --- | --- | --- | --- |
| 1. Child Age | - |  |  |  |  |
| 2. Child Nocturnal Sleep Duration | $r = -.160^{**}$<br>CI = [-.23, -.07]<br>d = -.32 | - | | | |
| 3. Child Sleep Problems | $r = -.26^{**}$<br>CI = [-.34, -.18]<br>d = -.54 | $r = -.19^{**}$<br>CI = [-.27, -.11]<br>d = -.39 | - | | |
| 4. Parent Sleep Related Impairment | $r = .01$<br>CI = [-.07, .09]<br>d = -.03 | $r = -.20^{**}$<br>CI = [-.27, -.12]<br>d = -.40 | $r = .17^{**}$<br>CI = [.09, .25]<br>d = .35 | - | |
| 5. Parent Sleep Knowledge | $r = -.07$<br>CI = [-.15, .01]<br>d = -.15 | $r = .12^{**}$<br>CI = [.04, -.20]<br>d = .24 | $r = -.04$<br>CI = [-.12, .04]<br>d = .09 | $r = -.04$<br>CI = [-.12, .05]<br>d = -.07 | - |

$r$ = Pearson's $r$ ; CI = Confidence Interval; d = Cohen's $d$
$^{**}$ . Correlation is significant at the 0.01 level (2-tailed).

Subsequently, a Kruskal-Wallis test was conducted to establish whether there was a difference in scores on the outcome variables between the child primary diagnoses groups. There was a statistically significant difference between child primary diagnoses and their average sleep hours, *H*(7) = 15. 70, *p* = .028, *d* = .25. Significant differences between primary diagnosis groups and parent sleep related impairment scores, *H*(7) = 20.66, *p* = .004, *d* = .31 were significant. Pairwise comparisons with adjusted p-values showed that children diagnosed with SEMH and ADHD had a statistically significant difference in their parent sleep related impairment score, albeit with a small effect size (*p* =.027, *r* = −1.98, *d* = .20). Children diagnosed with ADHD had a mean rank score of 340.52 compared with 93.5 in children with SEMH. There were no statistically significant differences in scores between the other primary diagnosis groups. There was no significant difference between child primary diagnosis group and parent sleep knowledge scores, *H*(7) = 13.22, *p* = .067, *d* = .21. The results of the multiple regression analysis are displayed in Table 5.

**Table 5.** Moderated Multiple Regression Analysis.

| **Model 1** |  | **Parental Sleep Impairment** |
| --- | --- | --- |
| n = 582 |  | R = .184 |
|  |  | R<sup>2</sup> = .034 |
|  |  | F = 4.026 |
|  | B | p |
| Child Age | .154 | .161 |
| Child Primary Diagnosis | -.056 | .863 |
| Child Sleep Problems (centred) | .169 | **<.001** |
| Parental Knowledge (centred) | -.153 | .545 |
| Child Sleep Problems x Parental Knowledge | -.009 | .749 |
| **Model 2** |  | **Parental Sleep Impairment** |
| n = 582 |  | R = .221 |
|  |  | R<sup>2</sup> = .049 |
|  |  | F = 7.399 |
|  | B | p |
| Child Age | -.045 | .671 |
| Child Primary Diagnosis | -.012 | .969 |
| Child Nocturnal Sleep Duration (centred) | -.889 | **<.001** |
| Parental Knowledge (centred) | -.064 | .805 |
| Child Nocturnal Sleep Duration x Parental Knowledge | -.270 | **.017** |

(p < .05) associations are in boldface

In the first regression analysis, after controlling for child age and primary diagnosis, child sleep problems were a significant independent predictor of parental sleep related impairment when parental knowledge was a moderator (*p* = < .001). There was no evidence parental knowledge acted as a moderator in this relationship. In the second regression analysis, after controlling for child age and primary diagnosis, parental knowledge was not a significant independent predictor of parental sleep related impairment (*p* = .805). However, child nocturnal sleep duration was a significant independent predictor of parental sleep problems (*p* = .001). In the model, the interaction between parental sleep knowledge and child sleep problems was also statistically significant (< .05), suggesting that the main effects could be interpreted in relation to an interaction effect. Following the recommendation by Aiken *et al*. [39] a simple slope analysis was conducted to aid the interpretation for this interaction. Visual inspection of the slopes showed that there was a relationship between parental knowledge and parental sleep problems only when child sleep problems were at an average (*b* = −.66, 95% CI [-.98, −.33], t = −3.95, *p* < .001) or high level (*b* = −1.20, 95% CI [-1.68, −.73], t = −4.96, *p* < .001) compared with a low level (b = −.39, 95% CI [-.83, .05], t = −1.73, *p* = .085). Specifically, parents with average or high levels of sleep knowledge scores had lower parental sleep problems when child sleep problems were high compared to parents with lower levels of parental knowledge (Figure 1).

**Figure 1.**
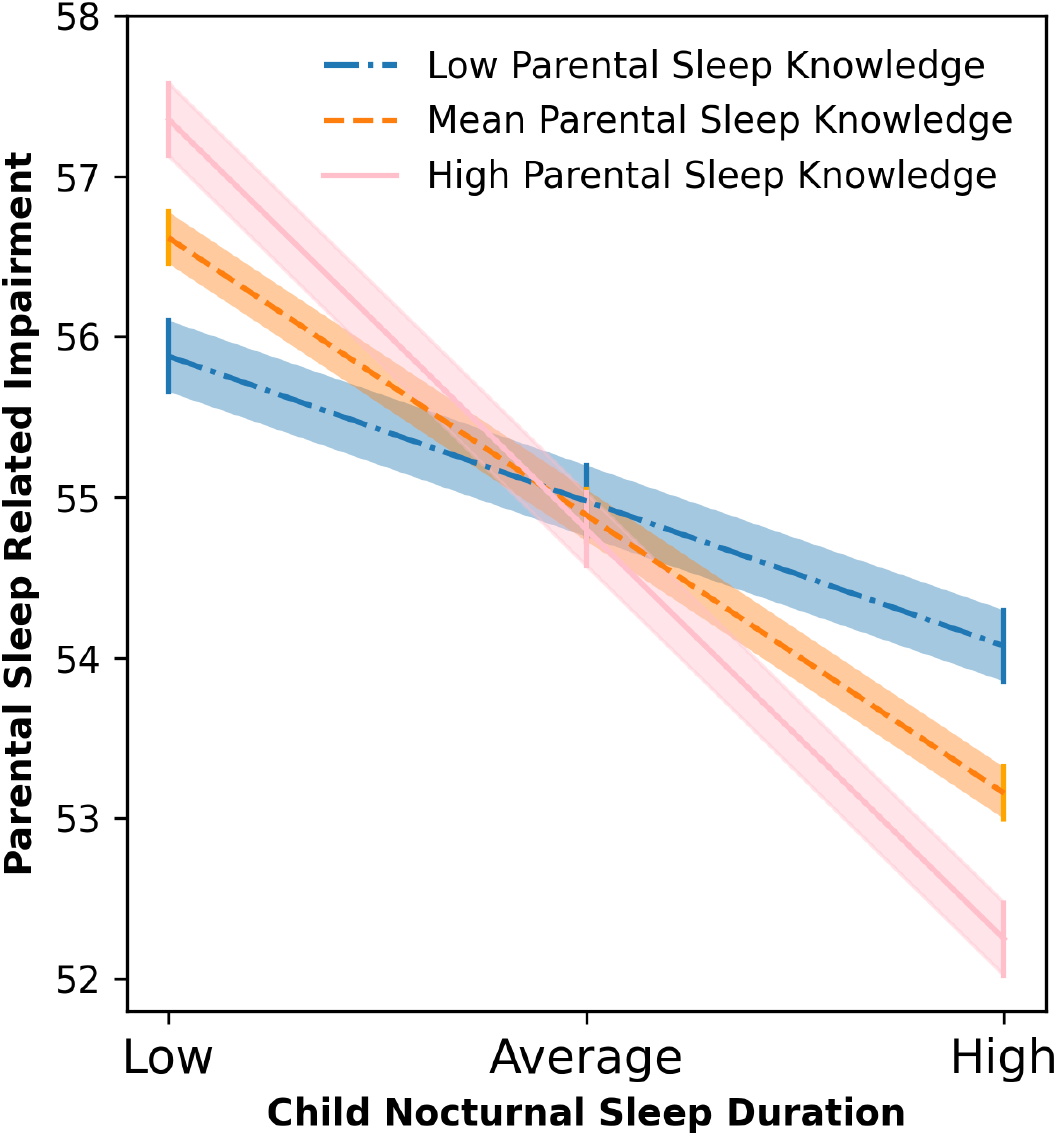
Moderation effect of parental knowledge on the relationship between total nocturnal sleep duration and parent sleep related impairment. Lines represent *Means* and bars and shades *Standard Errors*.

The percentage of parents answering each question correctly in the sleep knowledge questionnaire is shown in Table 6. Parents showed greater knowledge regarding bedtime routines (96% correctly answered question 6), and pre-bedtime behavioural practices (e.g. watching TV before bed). Parents appeared to have less knowledge regarding required duration of child sleep (67.9% parents incorrect) and recognising signs of a well-rested child (69.3% parents incorrect). Only 5.7% of parents answered all 8 questions correctly, with 14.1% answering half the questions correctly.

**Table 6.** Percentage of Parents Answering Sleep Knowledge Questions Correctly

| Question | % of Correct Answers |
| --- | --- |
| 1. Children who do not get enough sleep are more likely to be underweight than overweight; (False) | 64.9 |
| 2. Snoring indicates a child is sleeping well; (False) | 87.1 |
| 3. Being under- or overactive can be warning signs that a child is not getting enough sleep; (True) | 67.7 |
| 4. Watching TV in the bedroom makes it more difficult for children to fall asleep; (True) | 86.4 |
| 5. Children should have the same bedtime and wake time on weekdays and weekends; (True) | 78.4 |
| 6. Children only need a bedtime routine if they are having trouble falling asleep; (False) | 96.0 |
| 7. Well-rested children do not need an alarm clock to wake up in the morning; (True) | 30.8 |
| 8. The average school-aged child needs 8 hours of sleep per night; (False) | 32.1 |

## 4. Discussion

This study firstly aimed to establish a relationship between child sleep problems and nocturnal sleep duration with parental sleep impairment. Consistent with previous research, child sleep problems and nocturnal sleep duration were both significant independent predictors of parental sleep related impairment [40]. Secondly, our results demonstrated that parental sleep knowledge moderated the relationship between child nocturnal sleep duration and parental sleep related impairment. Specifically, increased parental knowledge resulted in longer sleep duration for children and reduced sleep disturbance for parents. Decreased parental knowledge was associated with shorter child sleep duration and increased parent sleep impairment. A significant overall association was identified between the child’s primary diagnoses and their nocturnal sleep duration. However, it should be noted that the correlation coefficients are modest, but significant likely due to the large sample size [41,42]. When comparing primary diagnosis, no significant differences were present in nocturnal sleep duration, suggesting children in our sample may experience a similar nocturnal sleep duration. Children diagnosed with ADHD had the lowest mean average sleep hours of 6.03, with the highest mean sleep hours of 7.58 experienced by children diagnosed with SEMH. Notably, both these durations are less than the recommended 8-11 hours for children aged between 2-17 (“How much sleep do children need?” 2017) [43].

Consistent with previous research, a significant association was found between child sleep problems and parental sleep impairment [9,10,40]. As child sleep problems and nocturnal sleep duration increased, parental sleep related impairment decreased. This finding was expected and provides further evidence for the impact of child sleep on parent sleep. Importantly, there may be a bidirectional relationship between child and parent sleep. Ng *et al*. [44] investigated whether maternal dysfunctional beliefs about sleep impacted their child’s sleep issues and found existence of an association. Conceivably, mothers of children with IDD who are exhausted and stressed due to their child sleep issues, experience elevated dysfunctional concerns regarding their own sleep. This may result in poorer sleep outcomes for these parents.

### 4.1. Parental Sleep Knowledge

Overall, parents demonstrated high sleep knowledge, considerably higher than studies conducted on parents with typically developing children [36,45]. Parents lacked knowledge on two specific topics, child sleep duration requirements, and the recognition of signs of a well-rested child. Encouragingly, 77.7% of parents answered over half the sleep knowledge questions correctly. This is in contrast to the findings of Schreck and Richdale [46] where less than 11% of parents of children aged between 2 and 17 years achieved this, suggesting sleep knowledge may have improved in recent years. There may be several reasons for this. Firstly, differing participant characteristics such as level of education could help to explain these varying findings [36]. Secondly, the consequences of insufficient sleep have generated greater media attention in recent years, often due to concerns over children’s increased screen time [47]. The media focus on this topic has possibly helped to increase parents overall general knowledge about sleep. This postulation is supported by the 86.4% of parents in this study who correctly answered the statement regarding children watching television in their bedrooms. Thirdly, our sample was parents of children with IDD who may have greater knowledge of sleep problems in their children as they relate to other health problems. Parents in our sample scored highly on two questions (snoring and bedtime routine) compared to previous literature. Many parents recognised snoring as an indicator of poor sleep, with 87.1% correctly answering this question compared to previous studies with parents of typically developing children [36]. However, Strocker and Shapiro [30] found that parents of children who had undergone a tonsillectomy demonstrated considerably more knowledge regarding snoring as a symptom of problematic sleep. In the present study, all participants have a child with a diagnosed IDD, many of which may experience increased hospital, doctor and medical interventions. This likely results in increased knowledge regarding their child’s condition.

Most parents in this study demonstrated strong knowledge regarding bedtime routines for their children. A large proportion, 78.4%, correctly acknowledged that children should wake and go to bed at the same time on weekends and weekdays. The vast majority of parents (96%), correctly marked the statement, ‘*children only need a bedtime routine if they are having trouble falling asleep*,’ as false. It seems that parents tend to understand the importance of bedtime routines for their children, a key recommendation of the [43,48]. Whether parents are able to enforce these bedtime routines will likely depend on the child’s disability, its severity and their social cognition ability Malow *et al*. [23].

The knowledge questionnaire revealed some knowledge gaps where parents scored lower than in other questions, highlighting specific areas where further education may be needed. The first of these were the signs of a well-rested child, with 69.3% of parents incorrectly marking, ‘*well-rested children do not need an alarm clock to wake up in the morning*,’ as false. In agreement, [24] established that parents lacked this specific sleep knowledge in their systematic review of literature. Secondly, only 32.1% of parents accurately answered the statement ‘*the average school-aged child needs 8 hours of sleep per night*.’ This is less than the recommended 9 hours of sleep or above per night for a school-aged child (“How much sleep do children need,” 2017) [43]. These findings were reflective of previous studies which found that parents underestimated their child’s sleep requirements [46,48,49].

### 4.2. Limitations and Future Directions

There are several considerations for this study. The first is that education level of parents and child medication was not included in the demographic questionnaire. This may have provided useful insights into the findings. For example, this study found that parents of children with ADHD had worse sleep quality than children with SEMH diagnosis based on the scores on CSHQ. It has been suggested that the use of stimulant medication often prescribed to children with ADHD can worsen sleep related issues [49]. In relation to this, there has been a 14.55% rise in ADHD medication use in children in Northern Europe between the years of 2001 and 2015 [50]. It could be speculated that increased use of medication worsened sleep difficulties of the children in the current sample. In addition, the differences in outcomes between child diagnoses may not have been clear due to uneven group sizes of diagnostic groups.

The study questionnaire was based on parental reports of their child’s sleep and is therefore subjective. Previous literature has identified differences between subjective and objective sleep reporting [51]. The use of objective sleep measurement would greatly improve the accuracy of the measurement of both the child’s sleep and parent’s sleep and provide further insight to this relationship.

Participants were predominantly recruited by convenience sampling via the social media accounts of UK charitable organizations that support families of children with neurodevelopmental conditions. Therefore, our sample is limited to parents/caregivers who engage in social media. In future studies, the collection of further demographic information of the families would contextualize the data and enable a more targeted approach in future research to include a more diverse sample.

Finally, it should also be noted that this questionnaire was not devised specifically for children with IDD. Although comparisons have been made with the previous literature in parents of typically developing children, there is little evidence to compare this questionnaire to other IDD families. It is currently unclear if parental sleep impairment is significantly more in this population compared to parents and families of typically developing children, and if this is related to the shorter sleep duration evidenced in children with IDD.

This study appears to be the first to investigate the role of parental sleep knowledge on child and parent sleep in a sample of children with IDD. This study provides evidence that efforts to improve parental sleep knowledge could reduce sleep problems in both children and parents. It also supports scholars who believe that educating parents should be the initial line of treatment [32]. Improving parental sleep knowledge, particularly in the areas where knowledge was lacking, i.e. sleep duration requirements of children and signs of a well-rested child should be a focus of future interventions. In addition, it is important to consider the impact on the whole family, both parents, siblings, and the child with IDD. Behavioural interventions for families in typically developing populations have previously provided positive outcomes [31,52,53] and have reportedly increased parents’ sense of control and coping ability [54]. A pilot randomized case-control trial with 20 parents of children with ADHD and 20 parents of typically developing children demonstrated that the training and monitoring of parents of children with ADHD, in regulating and supervising children’s sleep schedules, lead to positive changes in child emotion and behaviour [55]. The success of previous interventions coupled with the results from this study suggest that the implementation of educational based interventions to increase the sleep knowledge of parents could greatly benefit families of children with IDD experiencing sleep issues.

## Data Availability

Data are available on requests to the contact author

## Author Contributions

Conceptualization, E.J.H. and D.D.; methodology, E.J.H., A.J. and D.D.; formal analysis,E.J.H. and A.J.; data curation, E.J.H..; writing–original draft preparation, E.J.H. and A.J..; writing–review and editing, E.J.H., A.J., G.E. and D.D.; visualization, E.J.H and G.E.; funding acquisition, D.D.

## Acknowledgments

We would like to extend our appreciation to all the participants as well as to Giulio Gabrieli (Nanyang Technological University, Singapore) for his technical assistance.

## Abbreviations

The following abbreviations are used in this manuscript:

IDD: Intellectual and Developmental Disabilities
ASD: Autism Specturum Disorders
ADHD: Attention Deficit Hyperactivity Disorder
FASD: Fetal Alcohol Spectrum Disorders
SEMH: Social, Emotional, Mental Health

